# Chronic myelomonocytic leukemia with double activating *KRAS* mutations: molecular characterization of an uncommon case

**DOI:** 10.64898/2026.01.21.26344030

**Authors:** Enderson Murillo Ramos, Carlos Humberto Afanador Ayala, Katherine Andrea Palacio Rúa, Ángela Rodríguez Cárdenas, Gloria Cecilia Ramírez Gaviria, Claudia Marcela Cristancho Salgado, Nora Elena Durango Calle, Danna Cano Granda, Anyeli Hinestroza Córdoba, Juan Herrera Patiño, Carlos Mario Muñetón Peña, Gonzalo de Jesús Vásquez Palacio, Jair Fulgencio García Gómez, Vanessa Santiago Pacheco, Juan Sandoval Mesa

**Affiliations:** Laboratorio Integrado de Medicina Especializada (LIME), Hospital Alma Mater de Antioquia, Universidad de Antioquia, Medellín, Colombia; Grupo de Genética Médica, Facultad de Medicina, Universidad de Antioquia, Medellín, Colombia; Fundación Colombiana de Cancerología, Clínica Vida, Medellín, Antioquia, Colombia

**Keywords:** Chronic myelomonocytic leukemia, double *KRAS* mutations, RAS/MAPK pathway, Synergistic activation, Prognostic biomarkers

## Abstract

Chronic myelomonocytic leukemia (CMML) is a clonal myelodysplastic/myeloproliferative neoplasm characterized by persistent monocytosis and heterogeneous risk of progression to acute leukemia. Mutations within the RAS/MAPK signaling pathway, particularly involving KRAS, are linked to a proliferative disease phenotype and adverse prognosis. We report the first Colombian CMML case harboring two concurrent activating *KRAS* mutations (p.G12S and p.G13D). Both variants were detected at variant allele frequencies of 17% and 21% in a female patient in her late 50s presenting with marked leukocytosis, anemia, and thrombocytopenia. The coexistence of these mutations suggests synergistic hyperactivation of the RAS/MAPK pathway, likely driving clinical aggressiveness and therapeutic resistance. This case delineates a rare molecular subgroup within CMML and highlights the critical role of comprehensive genomic profiling to improve prognostic accuracy and inform precision medicine approaches.

## Introduction

Chronic myelomonocytic leukemia (CMML) is a clonal neoplasm of myelodysplastic/myeloproliferative nature, characterized by persistent monocytosis, myeloid dysplasia, and the absence of the Philadelphia chromosome or *BCR::ABL1* rearrangement. Its clinical course is heterogeneous, with a risk of progression to acute myeloid leukemia (AML) in approximately 30% of cases, and a median estimated survival of 20 to 32 months^1^.

The RAS/MAPK pathway regulates key cellular processes such as proliferation, differentiation, and survival. Mutations in genes within this pathway— including *KRAS, NRAS*, and *CBL*—occur in 25–30% of CMML cases and are associated with a proliferative phenotype, poor prognosis, and potential resistance to hypomethylating agents^2^.

Under physiological conditions, *KRAS* cycles between an active GTP-bound state (*KRAS*-GTP) and an inactive GDP-bound state (*KRAS*-GDP) through its intrinsic GTPase activity^3^. This activity can be accelerated by regulatory proteins known as GAPs, with neurofibromin (NF1) being one of the main negative modulators^4^. NF1 enhances the hydrolysis of GTP to GDP, thereby facilitating the inactivation of *KRAS* and maintaining proper control over proliferative signaling^3^.

Mutations in codons 12, 13, and 61 of *KRAS* inhibit GTP hydrolysis, thereby maintaining the protein in its active form and driving uncontrolled cell proliferation. While individual mutations in codons 12 and 13 are common in patients, the simultaneous occurrence of two activating mutations such as p.G12S and p.G13D in a single CMML patient, is extremely rare and has not been extensively documented in the literature. This absence of reports limits our clinical and molecular understanding of such cases ^5,6^. Notably, double *KRAS* mutations have been described in colorectal cancer ^7^, where they are associated with poor prognosis and diminished response to treatment, suggesting that similar effects may occur in CMML.

This article presents a clinical case of a Colombian patient with CMML harboring double *KRAS* mutations and explores the potential functional, biological, and clinical implications, with a focus on the possible synergistic effect leading to hyperactivation of the RAS/MAPK signaling pathway.

### Case report

A female patient in her late 50s was evaluated at a tertiary oncology center, presenting with involuntary weight loss of 4 kg, without fever, skin rashes, night sweats, or recent infectious symptoms. On physical examination, she was afebrile, with stable vital signs, no lymphadenopathy, and no active bleeding. Mild hypoperfusion and diffuse low-intensity abdominal pain were observed. Initial complete blood count revealed severe leukocytosis, moderate anemia, and severe thrombocytopenia. Bone marrow aspirate showed 35% monocytes and 29% promonocytes. Flow cytometry of the bone marrow identified a population of aberrant mature monocytes (83.53%) and a blast population comprising 14.61%. Immunohistochemistry (IHC) demonstrated CD34 positivity in 15% of blasts with a central lacunar distribution, which also expressed CD117. The immunophenotypic profile was consistent with CMML (Figure 1A-C).

**Figure 1.**
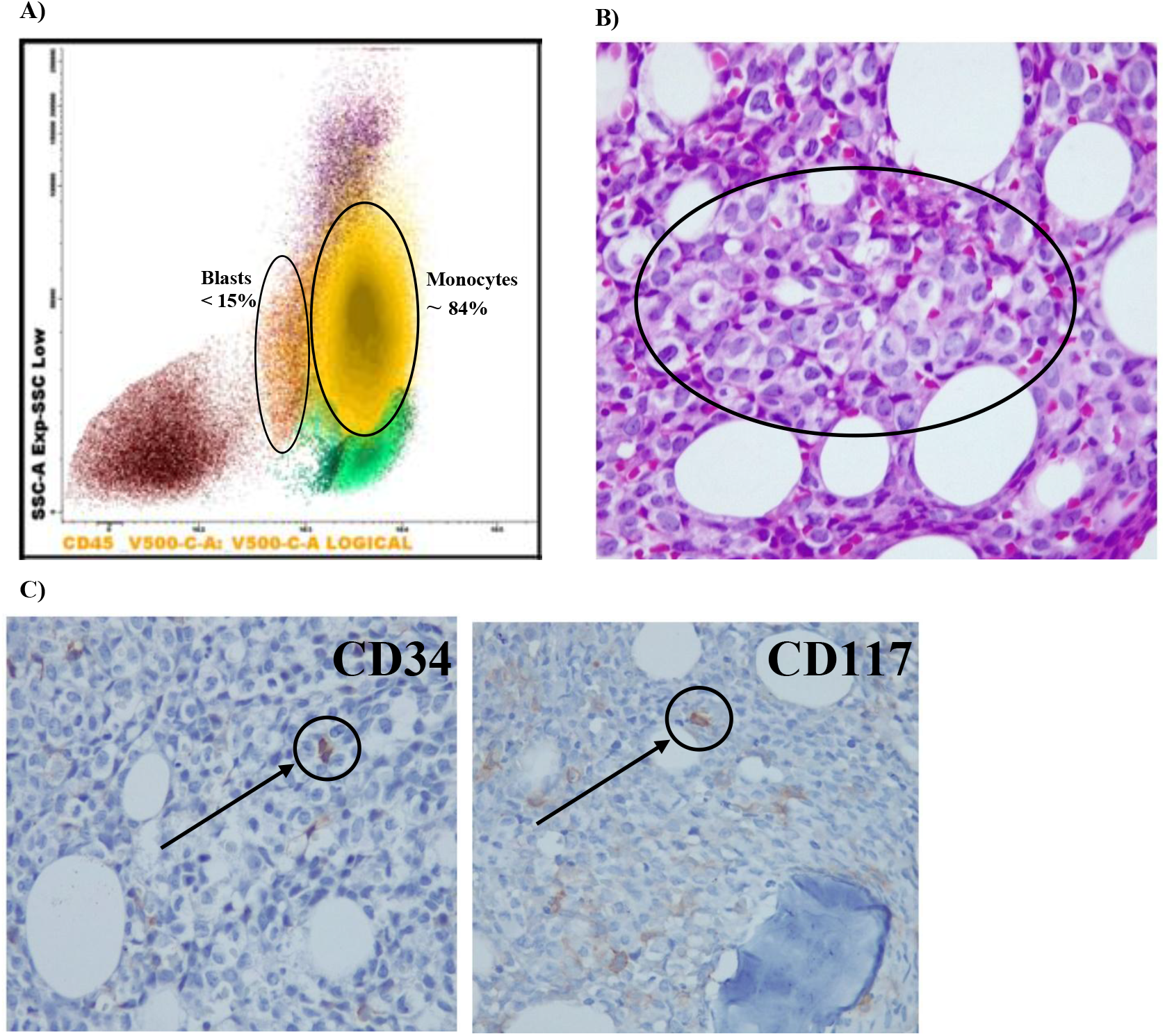
Findings compatible with CMML: morphology, flow cytometry, and immunohistochemistry. (A) Flow cytometry showed a relative mature monocytosis (83.53%), without expression of aberrant lineage markers (CD7−, CD56−), nor an increase in classical phenotype monocytes. A population of CD34+ blasts corresponding to 14.61% was identified. (B) Bone marrow biopsy stained with hematoxylin-eosin (40X), showing a hypercellular marrow (∼90%) with dysmature hematopoiesis and presence of myeloid and monocytic elements. (C) Immunohistochemistry (40X) for CD34 and CD117, with focal expression in less than 15% of cells, in an abnormal location. The ellipses in B and C highlight the areas of interest observed with monocytic infiltration and focal expression of CD34/CD117.These findings support a myelomonocytic entity with dysplasia, compatible with CMML.

Molecular analysis of bone marrow detected two activating heterozygous mutations in the *KRAS* gene: c.34G>A (p.G12S, rs121913530) and c.38G>A (p.G13D, rs112445441), with variant allele frequencies (VAF) of 17% and 21%, respectively, calculated using Mutation Surveyor® software version 5.2 (Figure 2A). Both variants have been previously reported independently in CMML and other hematologic neoplasms, and are classified as TIER I and oncogenic variants according to the AMP/ASCO/CAP criteria (Standards for the Classification of Pathogenicity of Somatic Variants in Cancer). Complementary molecular analyses did not reveal mutations in other genes frequently altered in CMML, including *SF3B1* (exons 14–16), *TP53* (exons 5–8), *JAK2* (p.V617F and exon 12), *calreticulin* (exon 9), *MPL* (exon 10), *NPM1* (variants A, B, D), *FLT3* (*FLT3*-ITD and TKD p.D835), *IDH1* and *IDH2* (exon 4), *NRAS* (codons 12, 13, 61), and *KIT* (exons 11 and 17).

**Figure 2.**
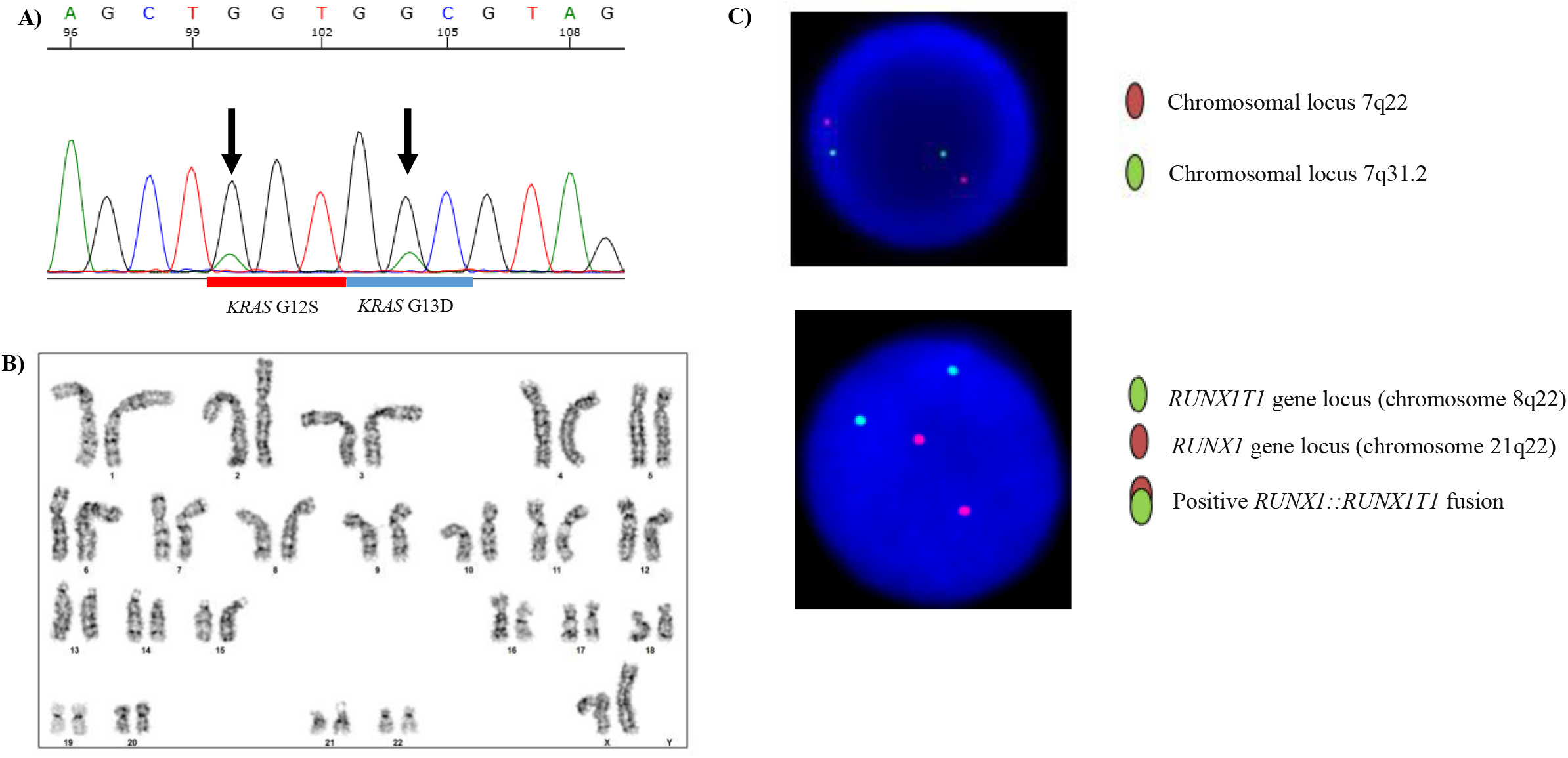
Molecular and cytogenomic studies (Sanger sequencing, karyotype, and FISH. (A) Chromatogram obtained by Sanger sequencing of exon 2 of the *KRAS* gene, which includes codons 12 and 13, where pathogenic variants c.34G>A (p.G12S) and c.38G>A (p.G13D) were identified. (B) Bone marrow cytogenetic analysis with a normal female karyotype: 46,XX, according to the ISCN nomenclature. (C) FISH showing no structural or numerical alterations in the long arm of chromosome 7, nor the presence of the translocation *t(8;21)(q22;q22)* associated with the *RUNX1::RUNX1T1* fusion.

The karyotype for leukemic states in bone marrow was normal (46,XX) in all analyzed metaphases (Figure 2B). Likewise, fluorescence in situ hybridization (FISH) analysis of bone marrow showed no gains, losses, or rearrangements in the regions PDGFRA (Locus 4q12), *PDGFRB* (Locus 5q32), Locus 7q22-q31.2, and *RUNX1::RUNX1T1* t(8;21)(q22;q22) (Figure 2C).

The patient began treatment with azacitidine 75 mg/m^2^/day subcutaneously for 7 days and symptomatic management with allopurinol. Bone marrow biopsy confirmed maturational changes and increased blasts (>15%) and the patient was classified as “fit” (ECOG 1, Karnofsky 90), considered a candidate for intensive treatment, including evaluation for allogeneic hematopoietic progenitor cell transplantation.

Currently, the patient is undergoing the third cycle of chemotherapy; she is asymptomatic, with preserved renal function and no transfusion requirements, although the previously documented neutropenia persists. She is hospitalized for oncologic-specific management. Her performance status is ECOG 2 (unable to perform paid work), documented weight loss is noted, with a current weight of 57 kg.

## Discussion

The RAS/MAPK signaling pathway is a central axis in the pathogenesis of CMML, with activating mutations in *KRAS* responsible for its constitutive activation, promoting cell proliferation and survival, and leading to oncogenesis^8^. In this report, we present the first described case of CMML with coexistence of two activating *KRAS* mutations—p.G12S and p.G13D—both located at codons 12 and 13 of *KRAS*, critical regions for protein inactivation. This composite pattern suggests a potential functional synergy that could enhance amplification of the oncogenic *KRAS* signaling pathway and lead to persistent hyperactivation of the RAS pathway, which may result in increased clinical aggressiveness and rapid disease progression, as well as resistance to targeted or conventional antineoplastic therapies^6,9^.

In solid tumors such as colorectal adenocarcinoma (CRC), concomitant *KRAS* mutations (p.G12D and p.G13D) have been described and are associated with rapid disease progression and chemotherapy resistance^7^. Although these are biologically distinct entities, constitutive *KRAS* activation represents a common molecular mechanism in oncogenesis. In the present reported case, the simultaneous presence of the p.G12S and p.G13D mutations could represent a phenomenon similar to CRC, with an adverse clinical prognosis for the patient

Mutations in *KRAS* directly affect the catalytic site, increasing the likelihood of *RAS* binding to GTP (active *KRAS*) and decreasing the interaction with regulatory proteins such as GAPs that promote hydrolysis and GDP binding, such as NF1 (inactive *KRAS*). This disrupts the protein’s activation-inactivation cycle and leads to prolonged signal transduction. In Figure 3, GTP hydrolysis rates are compared between wildtype *KRAS* (*KRAS*-WT) and relevant mutations in CMML, illustrating differential resistance to intrinsic and GAP mediated negative regulation^10,11^.

**Figure 3.**
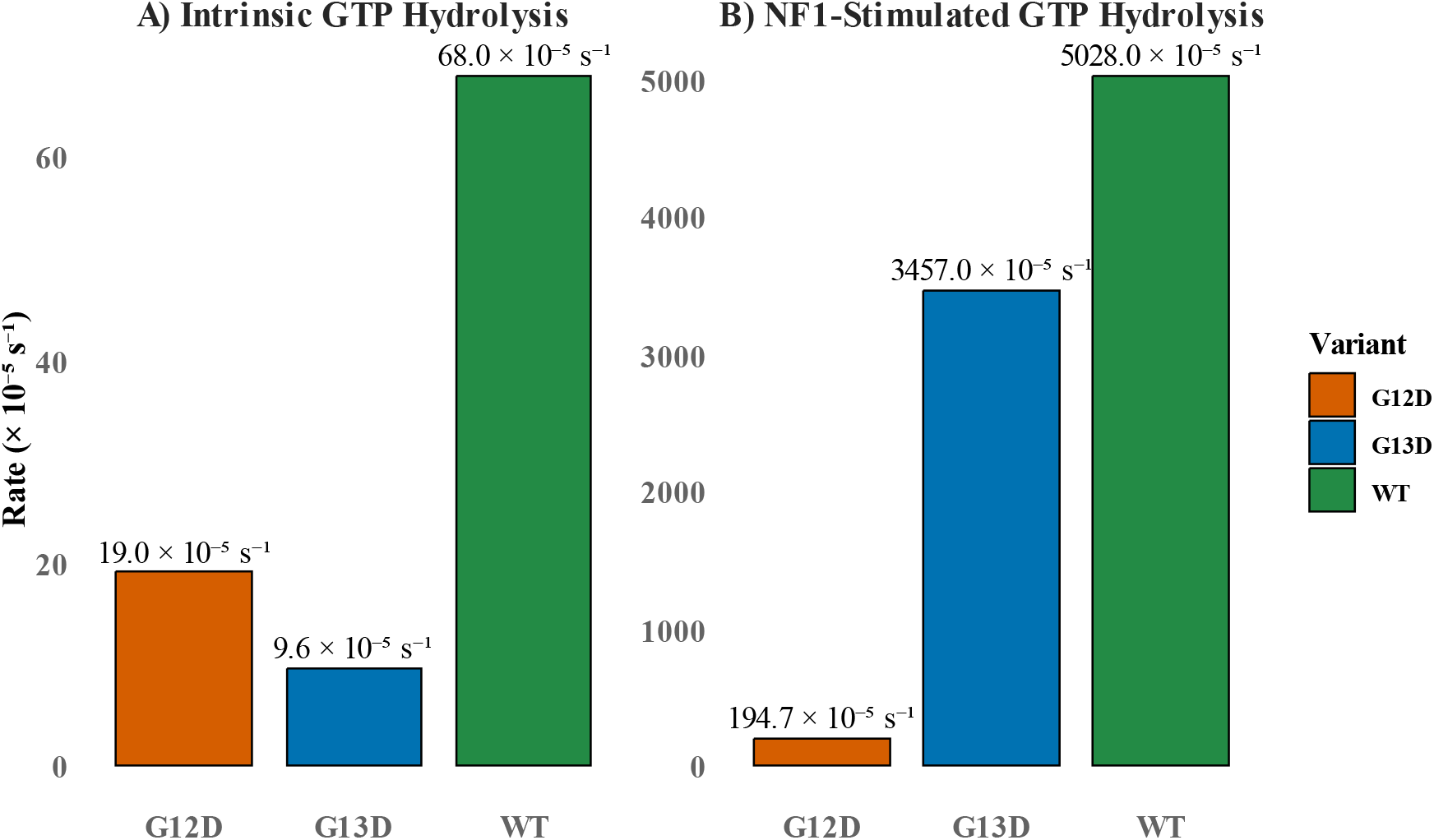
GTPase activity of *KRAS* in relevant mutational variants in the context of CMML, constructed from data published by Hunter^10^and Rajkumar^18^. (A) Intrinsic GTP hydrolysis by wild-type (WT) *KRAS* and oncogenic variants. The *KRAS*-WT protein (green bar) exhibits the highest basal GTPase activity (68 × 10^− 5^ s^− 1^), whereas the G12D mutant (orange bar) reduces this rate to 19 × 10^− 5^ s^− 1^, and G13D (blue bar) reduces it even further, to 9.6 × 10^− 5^ s^− 1^. (B) GTP hydrolysis stimulated by NF1, a physiological GAP protein that accelerates the conversion of *KRAS*-GTP to *KRAS*-GDP. In the presence of NF1, *KRAS*-WT (green bar) reaches a rate of 5.028 × 10^− 5^ s^− 1^, G13D (blue bar) maintains a partial response capacity with 3.457 × 10^− 5^ s^− 1^, while G12D (orange bar) shows marked resistance with a reduced rate of only 194.7 × 10^− 5^ s^− 1^. These results demonstrate that G12D is a strongly activating mutation resistant to negative regulation, whereas G13D retains partial sensitivity to GAP-mediated control. Together, this suggests that the coexistence of both mutations, as in the reported case of CMML, could induce constitutive and potentially synergistic *RAS*/ *MAPK* signaling pathway activation.

Similarly, other mutations at codons 12 and 13 significantly affect the regulatory mechanisms of *KRAS*; G12S is a driver mutation with a transformative potential similar to p.G12C, exhibiting decreased intrinsic GTP hydrolysis and resistance to regulation by *NF1*^12^. In contrast, the G13D mutation retains some sensitivity to GAPs but maintains elevated basal signaling of the MAPK pathway (Figure 3) ^13,14^. Therefore, the “double hit” (double mutation) presented in this clinical report could result in persistent activation and deregulation of the *RAS* pathway. Functional studies demonstrate that G12X and G13X variants, such as G12D, G12S, and G13D, activate partially distinct signaling pathways. The simultaneous presence of these pathogenic variants may broaden the spectrum of activated routes, intensifying oncogenic signaling. Proteomic analyses have shown tyrosine phosphorylation-based signaling profiles that differ between G12D and G13D, suggesting possible functional cooperation between both mutations^13^.

Moreover, it is important to consider that the potential synergistic effects of these mutations might be determined not only by functional alterations in the signaling pathway but also by their allelic configuration. Although in our case it was not possible to establish whether the variants were in *cis* (same allele) or in *trans* (different alleles), both configurations are relevant; the *cis* arrangement may enhance signaling by acting on a single *KRAS* molecule, whereas *trans* could suggest functional convergence between distinct subclones, implying greater tumor heterogeneity. The coexistence of these mutations may result from selective pressure on the RAS pathway, duplication of mutated alleles, focal hypermutation, or errors in DNA ^15^ repair mechanisms that promote tumor genetic instability

In terms of clonal dynamics, the analysis of the observed VAF in this patient suggests that these variants are not subclonal but rather affect a significant proportion of the tumor population. VAF values above the typical subclone detection threshold (<10%) usually correspond to early-acquired events or dominant clones with direct functional relevance in tumor biology^1,2^. In this context, the presence of both mutations with intermediate VAFs could suggest significant clonal expansion and direct involvement in the aggressive phenotype observed in this patient^3,5,16^.

Clinically, this dual activation may explain the rapid disease progression and the high blast burden observed in the bone marrow. *RAS* mutations are associated with reduced response to *FLT3* inhibitors and treatments based on azacitidine and venetoclax, both in CMML and other myeloid neoplasms^3,17^. Multicenter studies correlate these alterations with lower overall survival, increased risk of transformation to acute myeloid leukemia, and therapy-refractory disease^16^.

Given the high-risk biological profile, these patients are candidates for allogeneic hematopoietic progenitor cell transplantation (allo-SCT) from early clinical stages. In the present case, the patient meets the criteria established by international guidelines; although evidence regarding the specific impact of *KRAS* mutations on post-transplant outcomes is still limited, a higher relapse rate has been observed in patients with mutations in the RAS pathway, indicating the need for continuous molecular monitoring after transplantation^19^.

The incorporation of additional molecular analysis tools, such as next-generation sequencing (NGS) and single-cell sequencing, is essential to improve molecular classification, prognosis, early detection of molecular relapse, and to guide effective therapeutic decision-making. Furthermore, it is crucial to expand the functional and expression characterization of these dual mutations in myeloid models and validate them in larger clinical cohorts, with the aim of developing personalized therapeutic strategies for this high-risk patient subgroup to improve survival rates.

## Conclusions

This is the first reported case in a Colombian patient with CMML showing the coexistence of two activating mutations in the *KRAS* gene (p.G12S and p.G13D), defining a rare molecular subgroup characterized by potential synergistic activation of the RAS/MAPK signaling cascade. This molecular profile is potentially correlated with an aggressive clinical phenotype and therapeutic resistance. These findings underscore the imperative for early molecular subgroup identification, refinement of genomic risk stratification models, and implementation of personalized therapeutic interventions, including early allogeneic hematopoietic stem cell transplantation or targeted therapies against the RAS/MAPK pathway.

## Acknowledgments

We acknowledge the LIME Laboratory at the Universidad de Antioquia for providing the facilities for the analyses and test development; the Fundación Colombiana de Cancerología Clínica Vida for supplying the patient’s clinical history; and Carolina Montoya Ruiz, PhD, for her valuable assistance in reviewing and refining the translation of the clinical case.

## Authors’ Contributions

## Data Availability

All data generated or analyzed during this study are included in the submitted article.

## Ethics Considerations

Written informed consent was obtained from the patient prior to the publication of this case report. Ethical approval for this study was granted by the Institutional Research Technical Committee (Comité Técnico de Investigación) of Fundación Colombiana de Cancerología Clínica Vida, Medellín, Colombia (Approval Act: Acta CTI AE 055 RC). The committee reviewed the study protocol and approved the use of clinical and molecular data for research and publication purposes

## Consent to Participate

Written consent for publication was obtained from both the patient and Fundación Colombiana de Cancerología Clínica Vida prior to the submission of this case report.

## Consent for publication

All authors are in agreement with the content of the manuscript.

